# Integrating measles wastewater and clinical whole-genome sequencing enables high-resolution tracking of virus evolution and transmission

**DOI:** 10.64898/2026.02.21.26346782

**Authors:** Sipho Gwala, Joshua I. Levy, Victor Mabasa, Kathleen Subramoney, Nkosenhle L. Ndlovu, Chris Kent, Maryam Ahmadi Jeshvaghane, Praneeth Gangavarapu, Menzi Sikakane, Natasha Singh, Mantshali Motloung, Lethabo Monametsi, Lebohang Rabotapi, Emmanuel Phalane, Mokgaetji Macheke, Fiona Els, Chenoa Sankar, Tshepo Motsamai, Sibonginkosi Maposa, Nishi Prabdial-Sing, Joshua Quick, Kristian G. Andersen, Kerrigan McCarthy, Mukhlid Yousif

**Affiliations:** Centre for Vaccines & Immunology, National Institute for Communicable Diseases, A Division of the National Health Laboratory Service, Johannesburg, South Africa; Department of Translational Medicine, The Scripps Research Institute, La Jolla, CA, United States; School of Biosciences, Institute of Microbiology and Infection, University of Birmingham, Birmingham, B15 2TT, United Kingdom; School of Public Health, Faculty of Health Sciences, University of the Witwatersrand, Johannesburg, South Africa; Gauteng City-Region Observatory (GCRO), a Partnership of the University of Johannesburg, the University of the Witwatersrand, the Gauteng Provincial Government and Organised Local Government in Gauteng (SALGA), Johannesburg, Gauteng, South Africa; Department of Virology, School of Pathology, Faculty of Health Sciences, University of the Witwatersrand, Johannesburg, South Africa

## Abstract

Measles outbreaks have surged globally in recent years, but current surveillance systems have limited capacity to monitor measles virus (MeV) transmission and evolution at population scale. Although MeV can be detected in wastewater, the public health potential of wastewater genomic surveillance for MeV remains largely unexplored. Here, we deploy sensitive, low-cost MeV wastewater genomic surveillance combining virus concentration, whole-genome amplicon sequencing, and bioinformatic analysis alongside routine clinical genomic surveillance during the 2024-25 outbreak in South Africa. Integrated phylogenetic analyses of wastewater and clinical MeV genomes revealed previously undetected interprovincial spread and transmission links not captured by standard N450 sequencing. Our findings demonstrate that wastewater-integrated whole-genome surveillance expands the coverage and resolution of routine MeV monitoring and provides a scalable tool to advance measles control and elimination efforts.

## INTRODUCTION

Measles, caused by the measles virus (MeV), is among the most contagious diseases, requiring >95% vaccination coverage for herd-based immunity^1^. Despite major reduction in global incidence after public vaccine introduction in 1963^2^, outbreaks have resurged following decreases in vaccination coverage^3,4^ due to ongoing health system weaknesses, vaccine hesitancy, disinformation, and the COVID-19 pandemic^5–7^. Surveillance programmes that identify areas of MeV transmission and immunity gaps^8^ are therefore critical to achieve the WHO Measles and Rubella Strategic Framework 2021–2030 vision of “a world free from measles and rubella”^9^. Meeting this goal will require surveillance strategies that are more comprehensive and sustainable, and build on existing measles programmatic surveillance tools including case-based surveillance, contact tracing and quarantine, and genome sequencing. Scalable approaches are required in both resource-rich and resource-limited contexts to provide timely, sensitive, and reliable data to inform public health responses.

During an infection, MeV is shed through respiratory secretions, oral fluids, and urine^10–14^ and can be detected downstream in wastewater^15–18^. As demonstrated during the COVID-19 pandemic^17,19–21^, wastewater and environmental surveillance (WES) can provide population-level insights into pathogen transmission with reduced sampling bias and serve as a valuable complement to clinical monitoring. Although WES has been shown to enable detection of wild-type MeV^15,17,22–25^, its use in MeV genomic surveillance remains limited. Genome sequencing of MeV in wastewater may improve tracking of genotype prevalence, help demonstrate chains of transmission, and identify new outbreaks ^15,26^.

Programmatic MeV sequencing conducted under Global Measles and Rubella Laboratory Network (GMRLN) guidelines targets a 450-nucleotide region at the 3’ end of the nucleoprotein gene (N450)^27–29^. Selected in 1998, when there were over 18 circulating MeV genotypes, the N450 region covers < 3% of the 15.9kb MeV genome and was designed to support genotype assignment and basic tracking of transmission pathways^27,30^. However, increased vaccination coverage and decreased measles transmission over the period 2000-2020 led to the elimination of nearly all but two circulating measles genotypes, B3 and D8^27^. As a result, N450 sequences do not provide sufficient resolution to track virus transmission^31,32^, making it difficult to differentiate between local and imported strains^27^. Recent advances in virus concentration, combined with tiled amplicon scheme design^33^, now enable low-cost whole-genome sequencing (WGS), including from wastewater samples with low viral loads and fragmented RNA ^33,34^.

In South Africa, measles-containing vaccine (MCV) is administered through the Expanded Programme on Immunisation (EPI-SA) at 6 and 12 months of age^35^, with supplementary mass immunization activities conducted every 3-4 years^36^. Vaccination coverage remains below the 95% herd immunity threshold^37^, and measles outbreaks occur intermittently^38^. Since August 2024, measles outbreaks have led to 931 and 2756 laboratory-confirmed cases in 2024^39^ and 2025^40^. These outbreaks coincided with a national rubella outbreak, beginning in late July (epidemiological week 31) 2024, and declining by December 31st, 2024 ^41^. Measles and rubella surveillance is conducted according to WHO guidelines, with fever-rash cases tested simultaneously for measles and rubella IgM antibodies. For a subset of suspected measles cases, throat swabs are collected for PCR testing and N450 sequencing following GMRLN guidelines. Both genotypes B3 and D8 have been detected in clusters of cases or outbreaks since 2021, and sequences are regularly submitted to the Measles Nucleotide Surveillance (MeaNS) database^42^.

Here, using combined wastewater and clinical MeV whole-genome sequencing, we demonstrate that wastewater-based MeV surveillance expands outbreak tracking and improves characterization of MeV spread and evolution. Through longitudinal multi-modal virus detection and sequencing across South Africa during concurrent measles and rubella outbreaks, we show that wastewater MeV positivity genotype prevalence closely tracks laboratory-confirmed fever-rash cases and reveals wild type virus spread not captured by available clinical surveillance. Wastewater-derived genomes cluster with clinical sequences from South Africa and identify transmission links and virus diversity that cannot be resolved using N450 sequencing. Together, these results establish wastewater genomic surveillance as a scalable approach for high-resolution monitoring of measles transmission and support its use in measles elimination efforts, including in resource limited settings.

## RESULTS

### Wastewater surveillance tracks community MeV burden over time

The 2024-2025 measles outbreak in South Africa affected all nine provinces, with over 50% of laboratory-confirmed cases reported in Gauteng province (Fig. 1a)^43^. To supplement fever-rash surveillance for measles, we previously expanded the South African national WES program to include MeV detection in week 10 of 2024^17^. To further improve MeV wastewater detection sensitivity, we compared effectiveness of new two virus concentration approaches, a single magnetic capture bead method (Dynabeads; method A) and a two-bead method (Dynabeads and Ceres Nanotrap; method B), with our initial method, centrifugal ultrafiltration (Centricon; method C) (**Supp. Fig. 1a**). While all three methods yielded similar viral loads on average (**Supp. Fig. 1b**), method A was the most time and cost-effective option (**Supp. Table 1**). Therefore, from week 41 of 2024 method C was replaced by method A for routine processing of wastewater samples.

**Figure 1:**
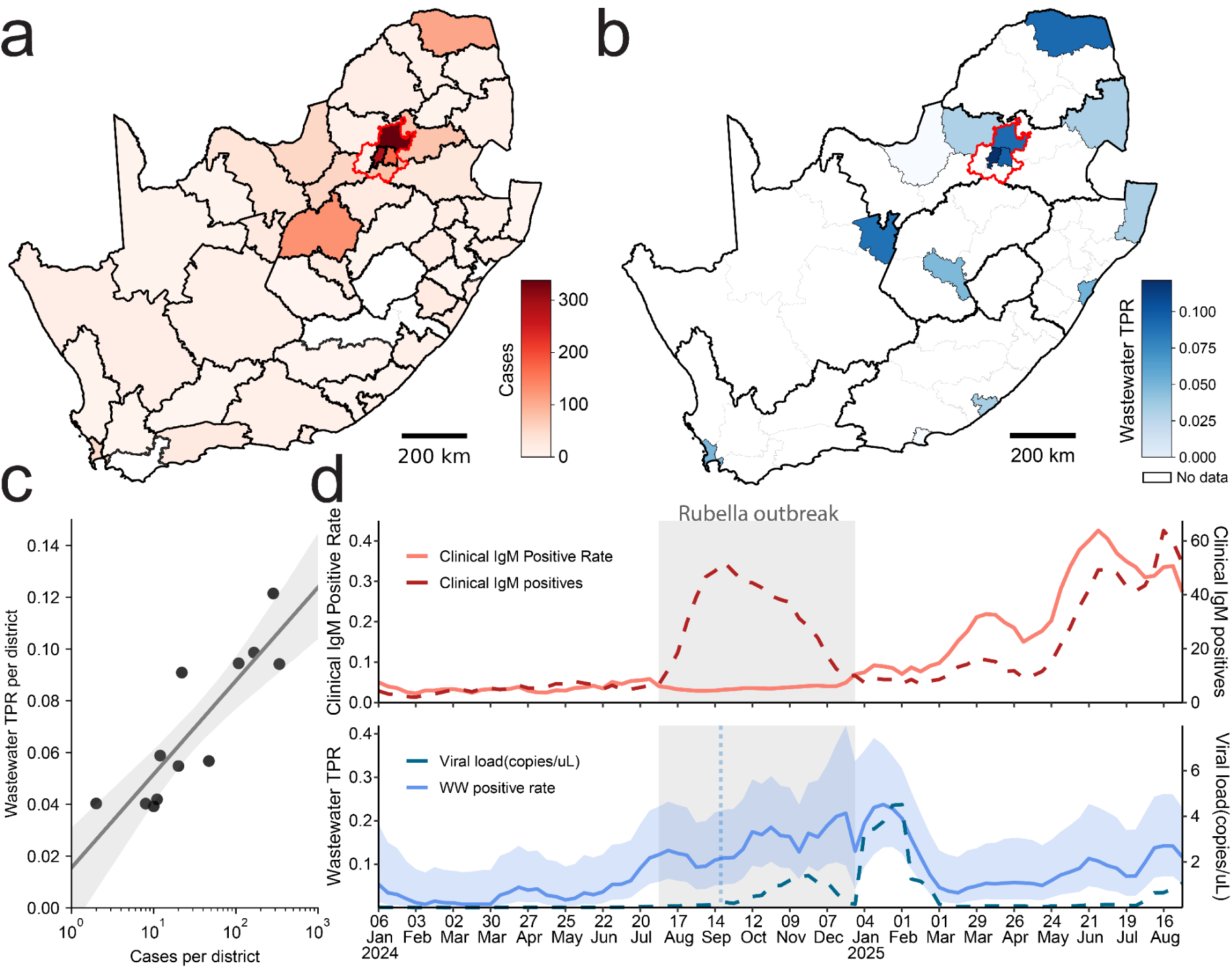
National wastewater MeV surveillance to supplement clinical monitoring in South Africa. For the period January 1, 2024 to August 31, 2025: a. Number of reported laboratory-confirmed measles cases by district. Gauteng province is indicated by red edges. b. Wastewater MeV test positivity rate (TPR) among national wastewater sites, aggregated by district. Districts where surveillance data was collected are outlined in black. c. District-level wastewater TPR versus cases per district, among districts with at least one wastewater detection, shown with best linear fit (R^2^=0.76). Bootstrap 95% confidence interval is shaded in grey. d. Aggregate national clinical positive cases (red dashed line) and clinical TPR (light red line) per epiweek from January 2024 through August 2025, smoothed with a rolling average of width 2 epiweeks (top). The period of increased rubella outbreak surveillance is shaded in grey. Wastewater TPR (light blue line) and 95% confidence interval (shaded) and aggregate wastewater viral load (blue dashed line) across all national wastewater sampling sites, with a vertical dotted line indicating the time of the change in virus concentration protocol. Tick marks are shown for the last day of every fourth epiweek.

From January 1, 2024 to August 31, 2025, we collected and analyzed 4,502 wastewater samples from 14 health districts across South Africa (**Supp. Fig. 2a**). We found that 395 samples (8.8%) were positive for MeV RNA. Wastewater test positivity rate (TPR) was highest in Johannesburg district (69/568, 12.1%), consistent with case reporting (Fig. 1b). Although we observed no detections in two districts with very low numbers of wastewater samples collected (n=21,24), wastewater TPR in all other districts ranged between 3.9-9.4%. Overall, we found that the wastewater TPR for each district was strongly correlated with the number of observed cases (Pearson’s r=0.87, p=2.2×10^−4^; Fig. 1c).

Wastewater sample counts were largely consistent over time (**Supp. Fig. 2a**), but clinical testing volume varied substantially, increasing from a baseline of ∼100 samples per week to over 1,000 per week between September and December 2024 on account of rubella outbreak surveillance efforts (**Supp. Fig. 2b**). As a result of the large volume of specimens submitted for rubella testing, measles clinical TPR remained low during 2024, despite substantial increases in both laboratory-confirmed measles cases and wastewater TPR (Fig. 1d). Comparing national wastewater TPR with clinical test results over time, we found that the number of clinical test positives and wastewater TPR were weakly correlated (Pearson’s r=0.32, p=2.3×10^−3^; **Supp. Fig. 2c**), but clinical TPR and wastewater TPR were not correlated (**Supp. Fig. 2d**). Additionally, we found that wastewater surveillance suggested an increase in TPR preceding the increase in measles cases in August-October 2024, although wastewater viral load did not increase until September 2024. These results indicate that wastewater is a strong correlate of laboratory-confirmed measles cases and can provide robustness to sampling related effects that may bias clinical surveillance results.

### Whole-genome wastewater sequencing recovers longitudinal genotype prevalence

Although clinical N450 sequencing volume increased in 2024 and 2025 relative to previous years, only 6.8% of laboratory-confirmed positive samples were sequenced, and uneven availability of sample collection and transport may limit the utility of genomic data to support outbreak response (Fig. 2a). To supplement tracking of MeV genotype prevalence using wastewater, we implemented a combined virus concentration and amplicon sequencing approach starting in October 2024 (**Supp. Fig. 3a**)^33^. Genome coverage had an approximately log-logistic relationship with viral load, with a mean expected genome coverage of ∼60% for samples with 10 copies/uL, and ∼95% coverage for a concentration of 100 copies/uL (**Supp. Fig. 3b**).

**Figure 2:**
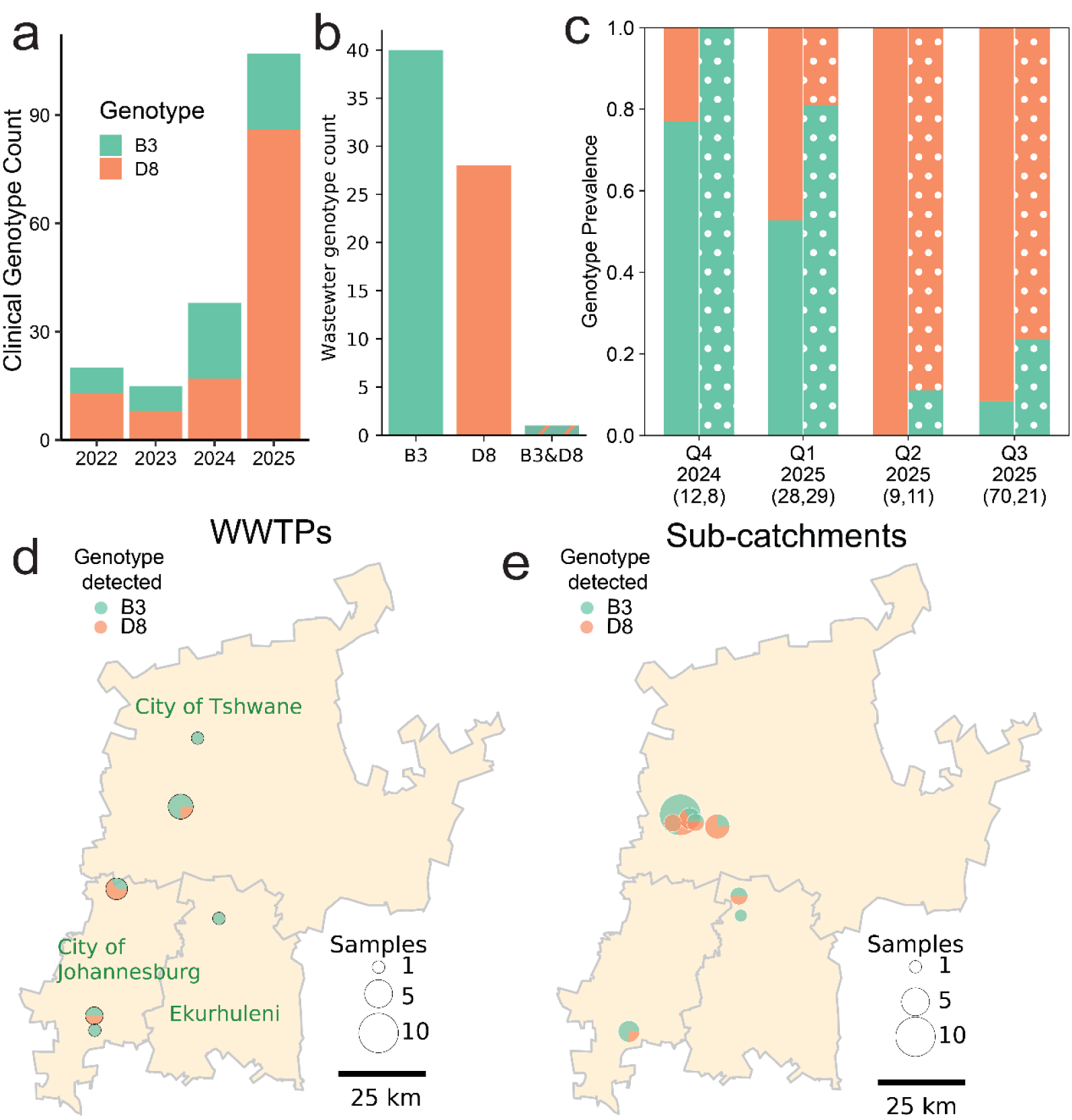
Wastewater amplicon sequencing recovers measles genotype dynamics. a. Counts of MeV sequences from 2022-2025 by year, separated by genotype, in South Africa. b. Estimated measles genotype composition for each wastewater sample using Freyja, across all sequenced samples. “B3” and “D8” indicate samples with estimated 100% composition of that genotype. “B3&D8” indicates that multiple genotypes are present in the sample. c. For the period October 1, 2024 to September 30th, 2025, quarterly clinical (solid bars) and wastewater (dotted bars) MeV genotype prevalence. d. Overall genotype prevalence for each national WWTP surveillance site in Gauteng. e. Overall genotype prevalence at each sub-catchment sampling site.

To estimate MeV genotype prevalence from virus mixtures, we extended our Freyja bioinformatic tool^44^ to incorporate available MeV genomes and cladistic nomenclature into genotype-defining mutation barcodes^45^. To test this sequencing and analysis workflow, we first simulated amplicon sequencing of genotype mixtures and analyzed genotype prevalence recovery (**Supp. Fig. 4a,b**). We found that our approach accurately recovered genotype prevalence for all reference samples including those with potential amplicon primer failure (R^2^= 0.99). To further validate our approach, we performed whole-genome sequencing of both vaccine strain (genotype A) MeV and clinical specimens from genotype B3 and D8. In each case, Freyja correctly assigned samples to their expected genotypes based on N450 sequencing (**Supp. Fig. 4c,d**). These results indicated that Freyja accurately recovered MeV genotype prevalence estimates for pure and mixed genotype samples in both simulated and real sequencing tests, respectively, comparable to previous results for SARS-CoV-2, MPXV, and ZIKV^34,44^.

To evaluate performance with real-world wastewater samples and compare results with clinical surveillance, we analyzed all available wastewater sequence data. Of the 395 samples that tested positive, 69 (17.5%) samples were sequenced, of which 51 had sufficient coverage to recover genotype prevalence estimates. Among these, 50 of them contained only one genotype (35 B3 and 15 D8), and one sample contained both B3 and D8 (Fig. 2b). We did not identify any samples with genotype A, indicating that MeV detected in wastewater derived from wild type infections rather than vaccine-derived shedding.

When grouped by quarter-year from Quarter 4 (Q4; Oct, Nov, Dec) 2024 to Q3 (through August) 2025, wastewater and clinical estimates of genotype prevalence were closely correlated (Cochran–Mantel–Haenszel X^2^ =13.3, p=2.6×10^−4^; Fig. 2c). Both surveillance demonstrated a consistent shift in genotype prevalence from predominantly genotype B3 detections in late 2024 to mostly D8 detections in Q2 and Q3 2025.

### Multi-scale wastewater sampling recovers fine-grained spatiotemporal dynamics

To study the geographic spread of MeV genotypes, we focused on wastewater samples from Gauteng province (88% of all wastewater sequences) and stratified into large-scale wastewater treatment plants (WWTPs) and “upstream” sub-catchment sites. At the WWTP level, 3 of 6 sites with positive detections had both B3 and D8 genotypes, while only B3 was detected at the other 3 sites (Fig. 2d). We then compared WWTP genotype detections with “upstream” sub-catchment sites in each district. In both City of Johannesburg and City of Tshwane districts, we observed both genotype B3 and D8 detections at WWTP and sub-catchment level. In Ekurhuleni, both genotypes were detected at the sub-catchment level, but only B3 was detected in district WWTPs (Fig. 2e).

To investigate the combined spatiotemporal dynamics of genotype spread, we further stratified by quarter-year, and compared across clinical (Fig. 3a) and wastewater surveillance (Fig. 3b,c). In Q4 2024, clinical surveillance identified genotypes B3 and D8, whereas wastewater detected only B3 at both the WWTP (Fig. 3b) and sub-catchment levels (Fig. 3c). D8 was first detected via wastewater in Q1 2025, when D8 detections for both wastewater and clinical surveillance were restricted to City of Tshwane and City of Johannesburg, but expanded to Ekurhuleni in Q2 2025. In Q2 2025, only genotype D8 was detected in both surveillance modes, and genotype B3 was again detected in both in Q3 2025. Together, these results suggest that MeV genotype introductions drive rapid geographic spread that can be tracked across spatial scales using wastewater.

**Figure 3:**
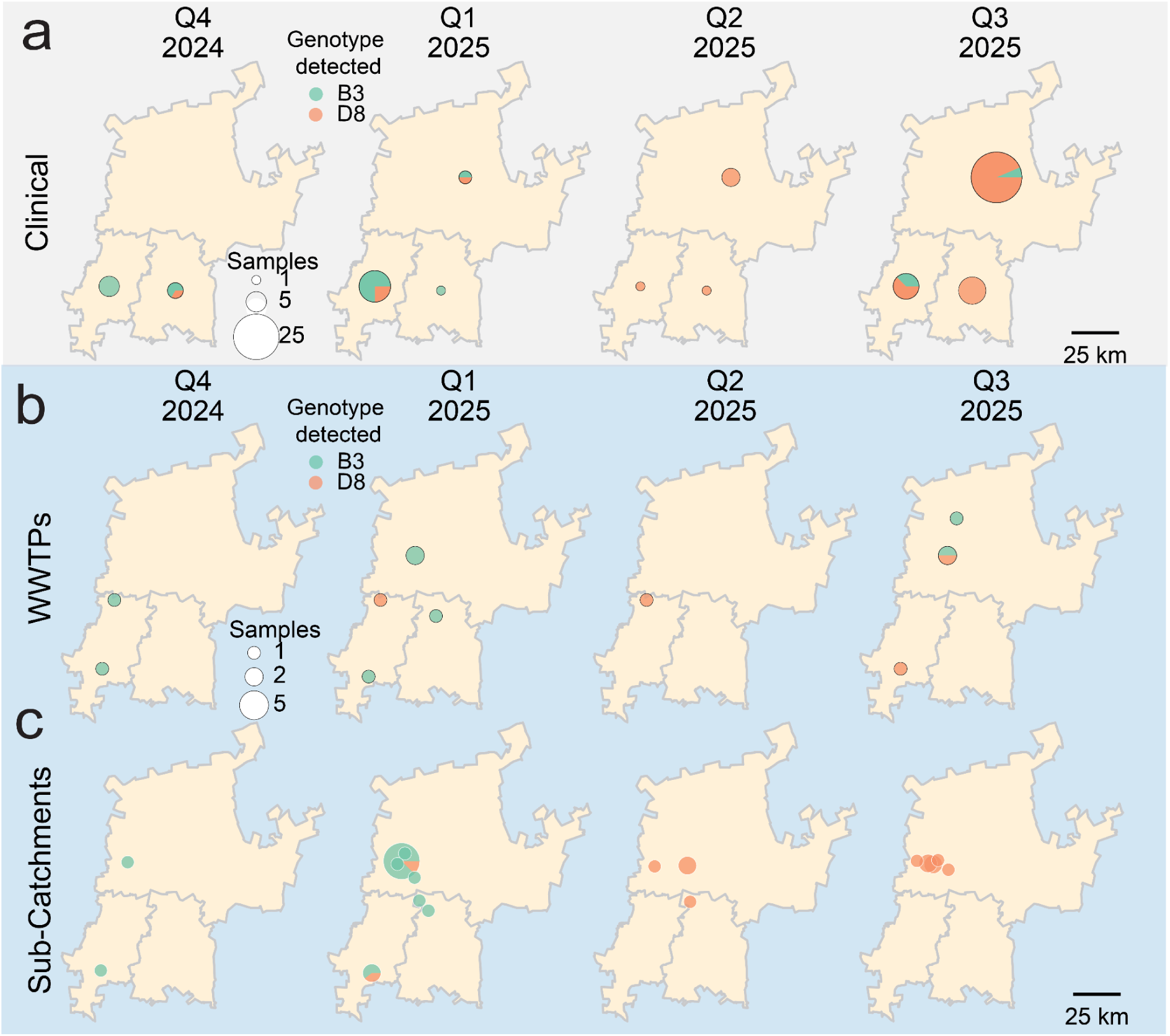
Wastewater recovers multi-scale geographic spread. For the period October 1, 2024 to September 30th, 2025, quarterly genotype prevalence across all clinical sequences (a), and wastewater sequencing from national WWTP sites (b), and sub-catchment sampling sites (c). Wastewater sites are shown at the geographic position of the site, with a small random perturbation added to sub-catchment site positions, while clinical genotype prevalence is shown centered about the district centroid.

### Minority variants reveal limited measles diversity in wastewater samples

Although Freyja genotype prevalence estimates indicated that nearly all samples were composed of a single MeV genotype, each genotype contains substantial virus diversity without corresponding cladistic designations. To characterize intra-genotype diversity present in wastewater, we next focused on minority single nucleotide variants (SNVs) in wastewater. We found a median of >600 consensus-level SNVs relative to the reference genome per sample, corresponding to highly dominant variants (here defined as >90% SNV frequency) among circulating MeV (**Supp. Fig. 5a**). However, we observed a median of only 10 non-dominant SNVs per sample (between 10 and 90% frequency; **Supp. Fig 5b**), indicating that circulating viruses have relatively little diversity and therefore support the use of simple consensus-based sequence generation.

To compare this with corresponding results in the N450 region, we performed the same analyses using only reads present in this part of the genome (**Supp. Fig. 5a,b**; right column). When restricted to the N450 region, we found that samples contained a median of only 20 highly-dominant SNVs, and 0 non-dominant SNVs, suggesting that N450 sequencing was unable to accurately capture circulating virus diversity.

### Wastewater-integrated genomic epidemiology recovers MeV evolution and spread

Because routine MeV sequencing has primarily targeted the N450 region, MeV sequence evolutionary dynamics in South Africa have remained underexplored. To enable complete genome analyses, we performed our whole-genome amplicon sequencing and bioinformatic workflow on 39 recently collected clinical samples. We then used a simple consensus calling approach with a 90% SNV frequency cutoff to recover virus sequences for both wastewater and clinical samples.

To analyze the relationships between clinical and wastewater sequences, we reconstructed integrated maximum likelihood phylogenetic trees for both genotype B3 and D8, combining newly generated genomes with all publicly available near-complete sequences (Supp. Data 1) to place wastewater and clinical viruses in a shared evolutionary context.

For genotype B3, we found that all clinical and wastewater sequences from South Africa were associated with a single clade (Fig. 4a) that includes sequences from recent outbreaks in the United States^46^ and the Netherlands (Fig. 4b). We observed that all clinical sequences from South Africa had descendants or sibling sequences observed in wastewater. However, some wastewater samples from Gauteng province clustered instead with clinical sequences from elsewhere in South Africa, with no related clinical sequences from Gauteng (Fig. 4b; black box). Multiple wastewater sequences in this cluster had no known clinical sequence ancestor, indicating that clinical sequencing may have missed a subset of recent outbreaks, or that these samples were not part of our retrospective sequencing.

**Figure 4:**
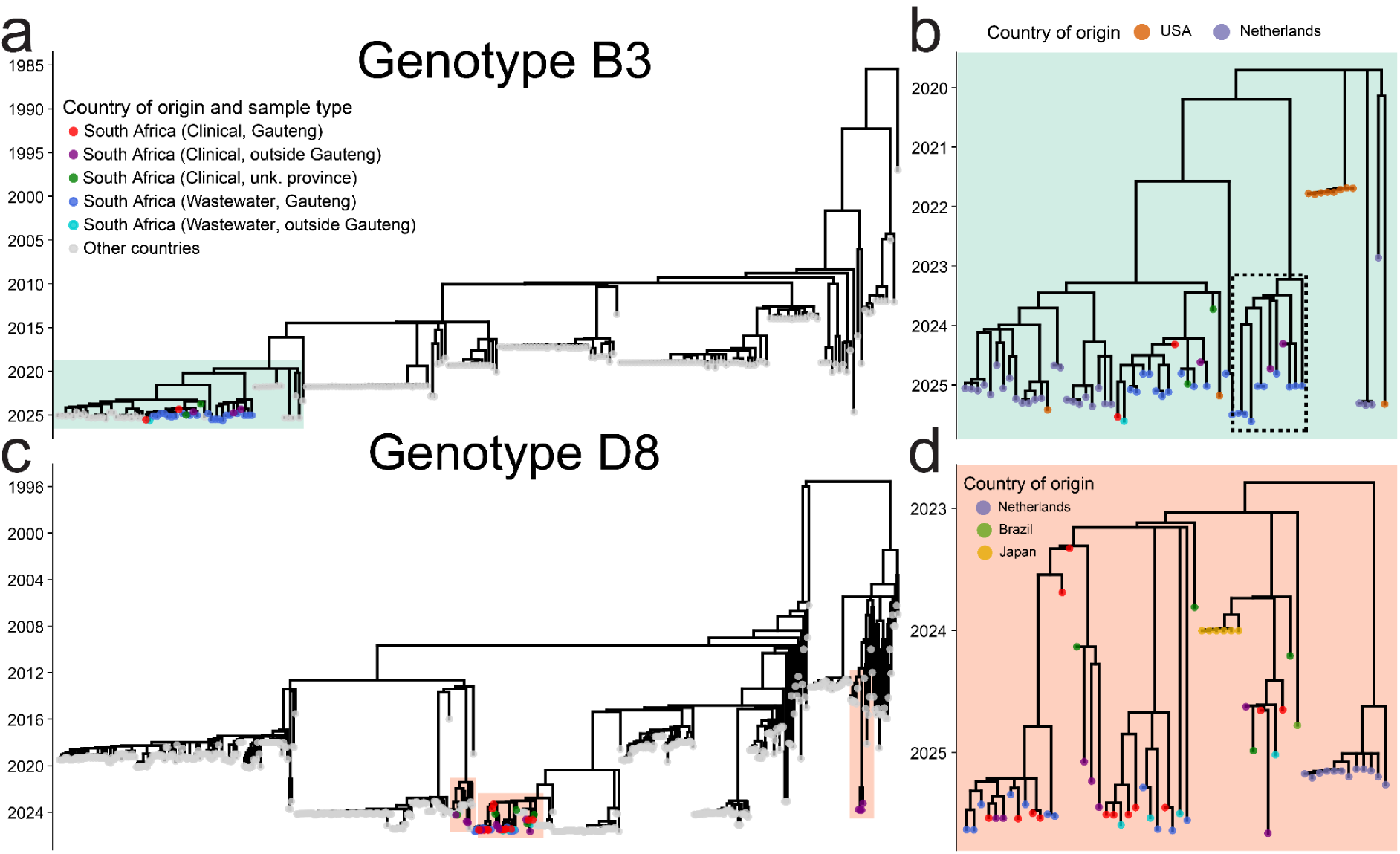
Wastewater genomic epidemiology identifies MeV evolution and spread. a. Time-calibrated maximum likelihood phylogeny for MeV genotype B3 whole-genome sequences. Inset region (teal rectangle) indicates a subtree containing sequences from South Africa. b. Subtree of wastewater and clinical sequences from South Africa and closely related whole-genome sequences. The dotted black box indicates wastewater sequences without a clinically-observed relative in Gauteng province. c. Time calibrated phylogeny for MeV genotype D8 whole-genome sequences, with inset regions (light orange rectangles) indicating South African sequences. d. Subtree of genotype D8 sequences containing wastewater and clinical case-derived sequences from South Africa.

For genotype D8, we found that South African sequences were associated with three distinct clades (Fig. 4c). While clinical sequences were found in all three clades, wastewater-derived sequences were only found in a clade with sequences derived from Japan, Brazil, and the Netherlands (Fig. 4d). In this clade, we again observed that most wastewater sequences descended from recent clinical sequences. Sequences from South Africa all clustered together, indicative of continued domestic spread rather than any clear MeV importation. Another cluster of South African sequences appeared to descend from two separate, but related outbreaks in Italy^47^ and Russia (**Supp. Fig. 6a**). The other D8 sequences from South Africa clustered tightly with one another, but have no recently sequenced relatives, with the most recent genome sequences from this cluster collected in 2014^48^ (**Supp. Fig. 6b**).

Finally, because consensus sequences can obscure co-circulating intra-genotype diversity, we examined non-dominant SNVs (between 10% and 90% frequency) in wastewater samples to identify virus diversity not captured by consensus genomes. For both B3 and D8, we found many non-dominant SNVs that matched mutations in South African consensus sequences of the same genotype, indicative of local MeV diversity (Supp**. Fig. 7a,b**). However, mutations from outside the dominant genotype in the sample were rare and restricted to a small subset of genomic positions, especially for genotype D8. To test the feasibility of leveraging this non-dominant SNV information we used the recently developed WEPP tool^49^, but it did not recover multiple haplotypes for any sample estimated to be composed of only one genotype by Freyja. Together, these findings suggest that most samples were dominated by a single circulating virus population with low diversity, and suggests non-dominant MeV haplotype reconstruction is not currently feasible.

### Whole-genome sequencing is necessary for accurate transmission tracking

To investigate the utility of using whole genome sequences instead of N450 sequences for transmission tracking, we re-analyzed all clinical and wastewater sequence data and masked all nucleotides outside of the N450 region (**Supp. Fig. 8**). Unlike for whole-genome analyses, we found that sequences from South Africa no longer formed tight clusters, and were instead interspersed with other sequence clusters from elsewhere in the world, both for genotype B3 and D8 sequences. To quantify differences in clustering by country in whole-genome vs N450 phylogenetic trees, we used a measure of trait-specific clustering, Parsimony Score (PS; number of trait switches required to traverse the tree)^50,51^. Using paired permutation testing for PS, we found that the whole-genome tree had significantly more clustering by country for both genotypes (B3: ΔPS=8,p=3.1 × 10^−2^; D8: ΔPS=16, p=3.4× 10^−3^). These findings indicated that geographic virus transmission dynamics were more accurately represented using the whole-genome approach.

To determine if whole-genome sequence trees were overall better representations of MeV sequence relationships, we then used an Approximately Unbiased (AU) test^52^ to tree likelihoods compared across topologies. We found the AU test rejected the N450 tree for both B3 and D8 (B3: p = 3.35 × 10^−81^, ΔlogL =7.7 × 10^3^; D8:p = 8.4 × 10^−5^, ΔlogL = 1.5 × 10^4^), indicating that whole-genome sequence trees provide better representations of relationships among sequences. Together, our results suggest that N450 sequences are insufficient to accurately characterize MeV geographic spread and evolutionary dynamics.

## DISCUSSION

In this study, we demonstrate that whole-genome sequencing from clinical and wastewater samples enables high-resolution and population-scale tracking of virus diversity, evolution, and spread. Deploying our approach in South Africa, we found wastewater-derived genotype prevalence closely mirrored clinical patterns while providing complementary spatial resolution from large-scale WWTPs to “upstream” sub-catchments. The consistently low within-sample diversity observed in wastewater samples supported simple consensus-based genome recovery and direct integration into phylogenetic analyses with clinical sequences. These analyses revealed geographically structured spread, uncovered interprovincial transmission, and multiple chains of domestic transmission lasting more than two years.

Effective wastewater surveillance requires concentration methods that align with both pathogen chemical properties^53^ and operational constraints. Filter-based approaches, in particular, have been widely used during the COVID-19 pandemic to concentrate SARS-CoV-2 from wastewater, but require large sample volumes and specialized centrifuges^54,55^. In contrast, bead-based concentration supports streamlined, high-throughput processing with reduced manual handling. Our findings show that magnetic bead-based methods provide reliable, scalable, and cost-effective MeV recovery, consistent with similar findings for SARS-CoV-2^56^.

Clinical measles case counts were only weakly correlated with wastewater TPR, likely reflecting increases in clinical testing volume during the contemporaneous rubella outbreak. Vaccination is unlikely to explain observed wastewater signals: genotype A was not detected using Freyja, and post-vaccination shedding has been shown to be many orders of magnitude lower than shedding during wild-type infection^57^. Although extended MeV RNA shedding can occur, MeV RNA titers typically drop over 100 fold by 30 days post-infection and reach undetectable levels after 70 days^14^, suggesting wastewater MeV signals primarily reflect recent wild-type infections.

Although outbreaks usually involve a single clade or genotype, reflecting transmission from a single viral introduction^58,59^, we observed multiple simultaneous outbreaks of both genotypes B3 and D8. All genotype B3 clinical and wastewater sequences formed a single clade that clustered with sequences from recent outbreaks in the United States and the Netherlands. Genotype D8 sequences were placed in three distinct outbreak clusters, including one containing both wastewater and clinical sequences from both within and outside of Gauteng, and closely related to sequences from recent outbreaks in Brazil and Japan. The other two clusters were entirely composed of clinical sequences from outside of Gauteng province, indicative of limited interprovincial transmission.

Although wastewater genotype prevalence estimates and spatial analyses in this study are based largely on samples from Gauteng, our analyses integrate with clinical sequencing of samples from across South Africa and with publicly available sequences. Our findings indicate sustained MeV transmission both within and across provinces in South Africa of both genotypes B3 and D8. We recover temporal and geographic spreading dynamics and provide a model for tracking both domestic and international MeV transmission. Our whole-genome sequencing approach outperforms the N450 standard for MeV sequencing, providing resolution needed to distinguish closely related viruses and accurately infer transmission patterns. Minority SNVs observed in wastewater, although relatively few, provide key signals of emerging transmission lineages. Iterative integration of wastewater and clinical sequences, together with new tools including WEPP ^49^ may enable haplotype reconstruction for non-dominant viruses present in wastewater.

After remarkable gains in measles vaccine coverage over the past 50 years, global progress in measles control has begun to stall, and in some settings, reverse^3,60^. Surveillance capacity is currently limited, particularly in low- and middle-income countries, but is critical to guide vaccination efforts and assess progress towards elimination targets^60^. Our results show that wastewater surveillance can complement clinical monitoring and extend MeV genomic surveillance to the population scale. As measles surges globally, integrating wastewater genomics into routine surveillance can strengthen outbreak detection, improve tracking of transmission, and support global measles elimination efforts.

## METHODS

### Wastewater sample collection

The NICD has an expanded network for WES built on the pre-existing infrastructures of the poliovirus environmental surveillance programme. The design and implementation of this network, including the details of the sentinel sites and the characteristics of the population served, have been described previously^61,62^. In brief, the network comprises 28 wastewater treatment plants (WWTPs) across all nine provinces of South Africa, along with 19 additional sentinel sampling sites located within the catchment of three major WWTPs serving metropolitan municipalities and a high-traffic international point of entry in Gauteng Province. The program routinely monitors vaccine-preventable infectious diseases, and for this study, we analysed 4,502 wastewater samples collected between January 1, 2024 to August 31, 2025 during the peak of measles outbreaks in South Africa (**Supp. Fig. 2c**). Confidence intervals for TPR (fraction of MeV-positive samples) were calculated using Wilson’s method for binomial sampled random variables.

### Virus concentration, enrichment, and nucleic acids extraction

Initial attempts to concentrate MeV from wastewater were performed on seven wastewater samples using Centricon Plus centrifugal filtration units (Merck, Germany), referred to as Method C in this work^62^, and have been routinely applied in our laboratory for WES testing of viral pathogens. However, preliminary evaluations indicated low nucleic acid recovery for MeV, prompting exploration of alternative enrichment strategies. Method A involved automated viral enrichment using the KingFisher Flex Purification System (Thermo Fisher Scientific, USA) with a 24-deep-well head. Briefly, 5 ml of wastewater and 100 µl of Dynabeads (Thermo Fisher Scientific, USA) were added to wells in Deep-Well Sample Plates, reserving one well per plate for an extraction negative control (5,000 µl nuclease-free water and 100 µl Dynabeads)^63^. Nucleic acids were extracted using the MagMAX Wastewater Ultra Nucleic Acid Isolation Kit (Thermo Fisher Scientific, USA) with minor modifications to the manufacturer’s instructions. A 100 µl volume of eluate was transferred to sealable 96-well plates for immediate downstream testing. Method B was trialled on a small subset of samples (n = 7) using a combination of Ceres Nanotrap Microbiome A Particles (Ceres Nanosciences, USA) and Dynabeads for virus enrichment, following a published protocol ^63^.

### Measles digital PCR detection and quantification

Extracted nucleic acids were subjected to RT-dPCR assay for the detection and quantification of MeV as described and validated by Ndlovu et al.¹³. Briefly, QIAcuity One (Qiagen, Germany) and 8.5k nanoplates were used. Reactions were prepared in a final reaction volume of 13.32μl containing 8μl of extracted nucleic acids, 3μl of 4× One-Step Advanced Probe Master Mix, 0.12μl of 100× One-Step Advanced RT-Mix, 1.6μl of Enhancer GC, and a 20× Primer-Probe mix: (MVN1139-F: 5’-TGGCATCTGAACTCGGTATCAC-3’; MVN1213R: 5’-TGTCCTCAGTAGTATGCATTGCAA-3’; probe: 5’-TAMRA-CCGAGGATGCAAGGCTTGTTTCAGA-BHQ1–3’) targeting the N-gene of all MeV strains. Thermal cycling was performed under the following conditions: reverse transcription at 50 °C for 40 min, RT enzyme inactivation at 95 °C for 2 min, followed by 45 cycles of denaturation at 95 °C for 5 sec, and a combined annealing and extension step at 60 °C for 60 sec. All tests included the Omzyta vaccine (MSD Pty. Ltd., South Africa) as the positive control, as well as the extraction negative and the non-template controls.

### Clinical specimen collection, RNA extraction, and RT-qPCR detection

As part of the routine fever rash surveillance conducted at the NICD, clinical specimens, including blood, urine, and throat swabs, were collected from individuals presenting with suspected measles infection at healthcare facilities across all South African provinces between 2023 and 2025 following WHO measles surveillance guidelines^64^. Specimens were initially screened for measles-specific IgM antibodies. Positive specimens were subsequently subjected to nucleic acid extraction using the QIAamp Viral RNA Mini Kit (Qiagen, Germany), followed by RT-qPCR screening^65^. PCR-positive specimens were sequenced by Sanger or next-generation sequencing (Illumina) methods targeting the N450 region of the MeV N gene for genotype determination. The remaining nucleic acids were stored at –80 °C for long-term storage. For the present study, a subset of remnant specimens or previously extracted nucleic acids (n = 39) was retrospectively retrieved. Extracted nucleic acids were then used for amplicon-based whole-genome sequencing.

### Whole-genome tailored amplicon sequencing

Nucleic acids from MeV PCR-positive clinical and wastewater samples were subjected to a previously published whole-genome amplification protocol validated for measles virus (MeV) in clinical specimens with slight modifications^33,66^. Complementary DNA (cDNA) was synthesised using SuperScript IV Reverse Transcriptase and random hexamers (Thermo Fisher Scientific), followed by amplification in two primer pools using Q5 High-Fidelity 2× Master Mix (New England Biolabs, USA). Each 25 µl PCR reaction contained 12.5 µl Q5 High-Fidelity 2× Master Mix, 10 µM of either Pool 1 or Pool 2 primers, 3 µl of input cDNA, and nuclease-free water. Thermocycling conditions consisted of polymerase activation at 98 °C for 30 s, followed by 40 cycles of 98 °C for 15 s and 65 °C for 5 min. Amplicons were visualised using a 4200 TapeStation instrument (Agilent Technologies, Germany), quantified with the Qubit dsDNA High Sensitivity (HS) Assay Kit on a Qubit fluorometer (Thermo Fisher Scientific), and purified using 1.8× volumes of Agencourt AMPure XP beads to remove excess dNTPs and primers. Paired-end libraries were prepared using the Illumina DNA Prep Kit (Illumina, USA) according to the manufacturer’s protocol. Normalised libraries were pooled and sequenced (2 × 150 bp) on the NextSeq 2000 platform (Illumina, USA).

### Amplicon primer scheme design and validation

We used a previously validated 400bp primer scheme, designed to target all MeV genotypes (https://labs.primalscheme.com/detail/artic-measles/400/v1.0.0)^33^. To ensure that this scheme was sufficiently robust, we performed a two-stage *in silico* validation process by simulating mixture sequence data and analyzing it with Freyja. We first selected a subset of high quality sequences from each known genotype with available whole-genome sequences (genotypes A,B3,D4,D5,D8,D9,H1), and identified amplicons that could potentially fail as a result of mutations in the primer regions. We then simulated sequencing of mixtures combining these representative sequences using Bygul (https://github.com/andersen-lab/Bygul), and estimated genotype and prevalence using Freyja^44^. For laboratory validation of the scheme at NICD, we also sequenced pure vaccine strain and clinical virus samples using the amplicon sequencing approach and again estimated genotype identity and prevalence with Freyja.

### Bioinformatic processing and genotype prevalence estimation

We mapped all wastewater and new clinical sequencing reads to the MeV reference genome (NC_001498.1) using minimap2 (version 2.30-r1287), and performed primer trimming using iVar (version 1.4.4), using a minimum read length of 80 nucleotides following trimming. Modeling of genome coverage versus viral load was performed using an iterative re-weighting least squares approach for robust logistic regression, followed by bootstrap resampling to estimate 95% confidence intervals. Quantification of within sample heterogeneity was performed using SNV frequency estimates provided in Freyja “variants” outputs. Genotype prevalence estimates were performed with Freyja, using the latest whole-genome barcode set (timestamped 2025-05-25) available directly through Freyja, and on Freyja-barcodes (https://github.com/andersen-lab/Freyja-barcodes/tree/main/MEASLESgenome). For statistical testing of the association between wastewater and clinical genotype measurements, we used a time-period stratified Cochran-Mantel-Haenszel test (for the one mixed genotype sample, the dominant genotype was used). For both wastewater and clinical samples, consensus sequence generation was performed using iVar “consensus”, using a minimum SNV frequency of 0.9 for variant calling. SNVs with frequencies between 0.1 and 0.9 were separately extracted for analysis of potential evolutionary steps and the extent of the circulation of corresponding viruses.

### Phylogenetic analysis combining wastewater and clinical genomes

All publicly available near-complete genome MeV sequences (>80% length of reference and >80% non-ambiguous bases) were obtained from Genbank and aligned using MAFFT (version 7.505). Using these background sequences, phylogenetic tree inference was performed using a general-time-reversible (GTR) model in IQ-TREE(version 2.2.0.3) to identify B3 and D8 subtrees and fix incorrect metadata. Sequences without host or organism fields populated were excluded from further analyses. The newly acquired B3 and D8 wastewater and clinical MeV sequences with minimum 25% genome coverage were then integrated into corresponding subtrees. For each genotype, new and background sequences were aligned using MAFFT, phylogenies were inferred with IQ-TREE, and then time-calibrated phylogenetic inference was performed using treetime (version 0.9.6) with a GTR molecular clock model. Extraction and plotting of genotype-specific subtrees and within-genotype subtrees was performed using baltic (version 0.3.0). For N450-only analyses, sequences were aligned to the MeV reference genome using minimap2 and gofasta(v1.2.3), and trimmed nucleotides outside of the N450 region, prior to phylogenetic tree inference using the same approach as for the whole-genome sequences. All accession numbers for background sequences used in phylogenetic analyses are provided in Supplementary Data 1.

### Phylogenetic comparisons across sequencing approaches

After trimming sequences from the whole-genome tree without N450 coverage, parsimony score analyses for phylogenetic trees were performed using the Fitch parsimony function available in the phangorn (version 2.12.1) R package. Association Index was calculated first using phangorn to extract node descendants, and the country trait for each sequence was used to calculate entropy for each node using a custom R script. Paired statistical testing was performed by permutation of tip labels (10,000 resamplings) for both whole-genome and N450 trees and calculating the percentile of the observed test statistics to the resampled distribution. Approximately Unbiased (AU) testing was performed using 10,000 bootstraps in IQ-TREE.

## DATA AVAILABILITY

All MeV wastewater raw sequencing data and clinical genome sequences produced are available under NCBI BioProject PRJNA1377662.

## CODE AVAILABILITY

Freyja is available via GitHub (https://github.com/andersen-lab/Freyja) under an open-source BSD-2-Clause license (https://doi.org/10.5281/zenodo.6585067, all versions). Freyja can be downloaded via bioconda (https://bioconda.github.io/recipes/freyja/README.html) and mutation barcodes for measles analyses are available via the Freyja and the Freyja-barcodes repository https://github.com/andersen-lab/Freyja-barcodes. Code for all analyses is available at https://github.com/NICD-Wastewater-Genomics/Measles-Genomic-Surveillance-Paper.

## ACKNOWLEDGMENTS

We thank the NICD Centre for Vaccines and Immunology and its employees for providing the measles clinical surveillance data. We also appreciate the municipal workers, sample collectors, and laboratory technicians, for their sample collection and processing efforts. Special thanks to Namhla Madikane for her administrative support and assistance with fieldwork. We also thank all members of the Modjadji wastewater surveillance initiative. This work was funded by the Gates Foundation (057213 to KGA, KM, MY; 049272 to KM, MY; 050051 to KM, MY), National Institutes of Health (5T32AI007244-38 to JIL; 3U19AI135995-03S2 to KGA; U19AI135995 to KGA; U01AI151812 to KGA, UL1TR002550 to KGA).

## ETHICS DECLARATIONS

KGA has received consulting fees for advising on SARS-CoV-2, variants and the COVID-19 pandemic. The other authors declare no competing interests. The protocol was reviewed and approved by the University of the Witwatersrand Human Research Ethics Committee (MM220904). The NICD conducts all routine clinical surveillance, including surveillance of notifiable medical conditions, in a protocol reviewed and approved by the University of the Witwatersrand Human Research Ethics Committee (HREC) M210752 and under the legal authority of the National Health Act (no. 61 of 2003).

## Notes

### Author Declarations

The protocol was reviewed and approved by the University of the Witwatersrand Human Research Ethics Committee (MM220904). The NICD conducts all routine clinical surveillance, including surveillance of notifiable medical conditions, in a protocol reviewed and approved by the University of the Witwatersrand Human Research Ethics Committee (HREC) M210752 and under the legal authority of the National Health Act (no. 61 of 2003).

