## Supplemental Table and Figures for "Integrating measles wastewater and clinical whole-genome sequencing enables high-resolution tracking of virus evolution and transmission"

| Method | Reagents Needed | Processing Time | Cost per sample | Required equipment | Scalable? |
|---|---|---|---|---|---|
| One magnetic capture bead (A) | MagMAX Wastewater Ultra Nucleic Acid Isolation Kit with Virus Enrichment | 2 hours | 14 USD | Dynamag-50, heating block, vortex and 1.5/2 mL tubes centrifuge for manual processing and KingFisher Flex/Apex for automation | Yes, for automation |
| Two magnetic capture beads (B) | Ceres Nanotrap Microbiome A Particles, MagMAX Wastewater Ultra Nucleic Acid Isolation Kit with Virus Enrichment | 3 hours | 25 USD | Dynamag-50, heating block, vortex and 1.5/2 mL tubes centrifuge for manual processing and KingFisher Flex/Apex for automation | Yes, for automation |
| Centrifugal Filter (C) | Centricon Plus Centrifugal Filter (100 kDa MWCO), MagMAX Wastewater Ultra Nucleic Acid Isolation Kit | 3 hours & 30 minutes | 55 USD | Refrigerated centrifuge with capacity for 250 mL tubes, vortex, 1.5/2 mL tubes centrifuge | Yes |

**Supplementary Table 1: Cost, time, equipment and scalability of virus concentration and extraction methods.** Processing time estimates assume a batch of 24 samples. Prices include reagent and consumable costs per sample, as of December 10th, 2025.

**Supplementary Data 1: Accession numbers of closely related background sequences**

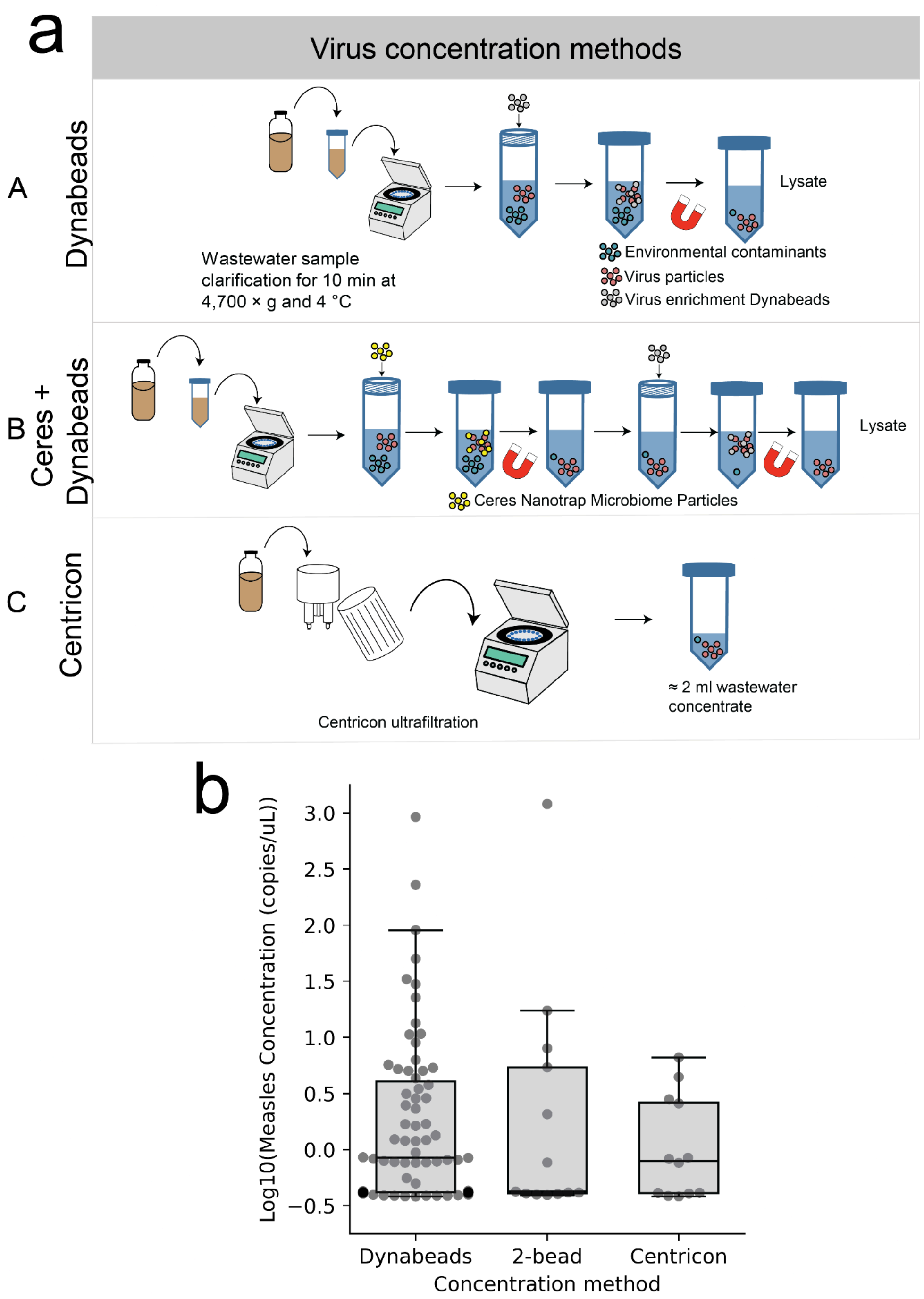


**Supplementary Figure 1: Validation of MeV sequencing workflow on control and clinical samples a.** Schematic of steps involved in each tested virus concentration method, including Dynabeads (Method A), Ceres NanoTrap Microbiome Particles and Dynabeads (Method B), and Centricon Ultrafiltration (Method C). **b.** Measles virus concentration for each of the tested virus enrichment approaches.

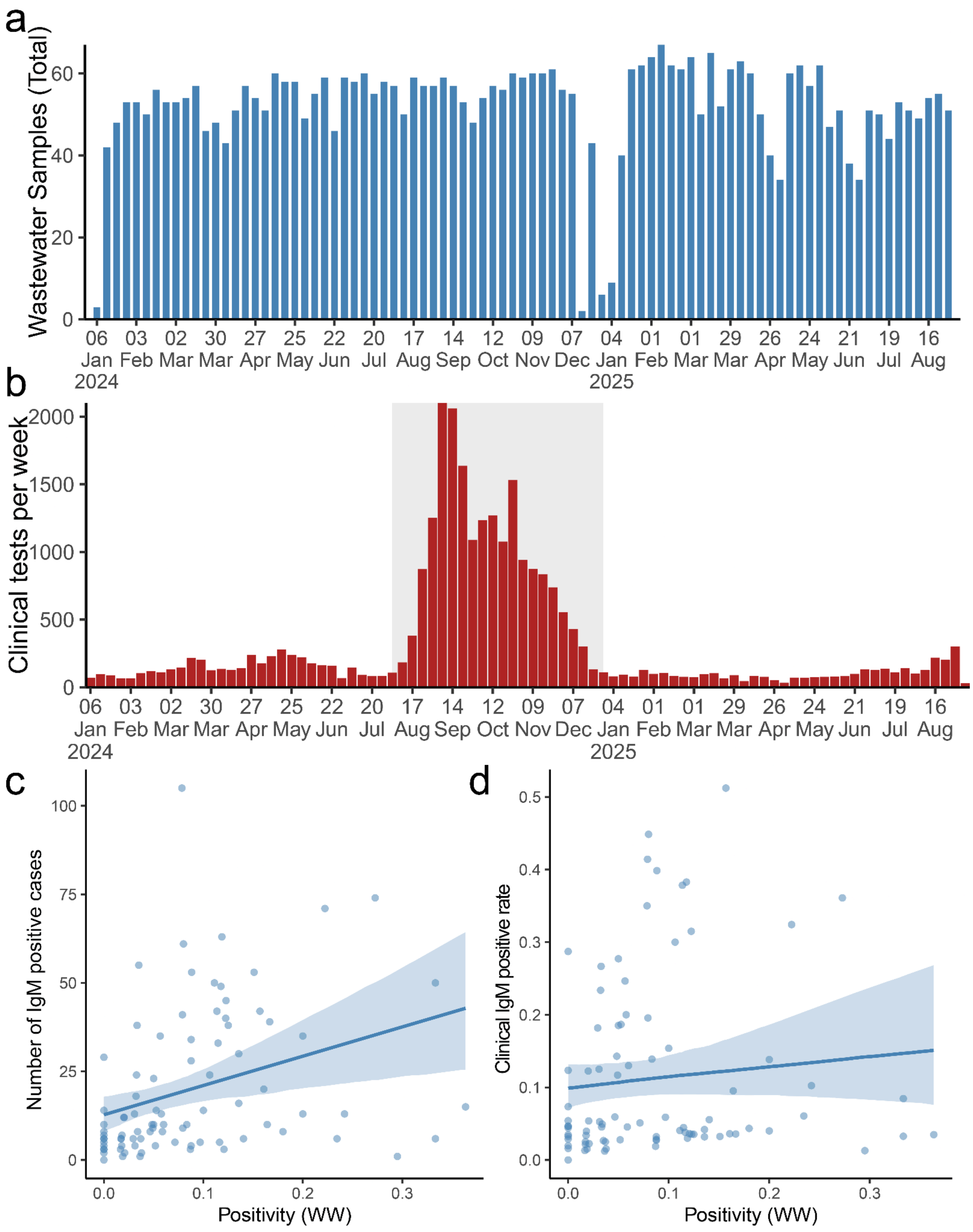


**Supplementary Figure 2: Wastewater sampling and clinical incidence correlation**
**a.** National wastewater samples tested by epidemiological week, with the last day of epiweek shown on the x axis. **b.** National clinical MeV tests performed by epidemiological week. The period of increased rubella outbreak surveillance is shaded in grey. **c.** Number of clinical IgM-positive tests from fever rash surveillance versus

wastewater positivity rate. **d.** National clinical IgM test positivity rate from fever rash surveillance versus wastewater positivity rate, with linear regression fit and 95% bootstrap confidence interval shown, using 1000 bootstraps.

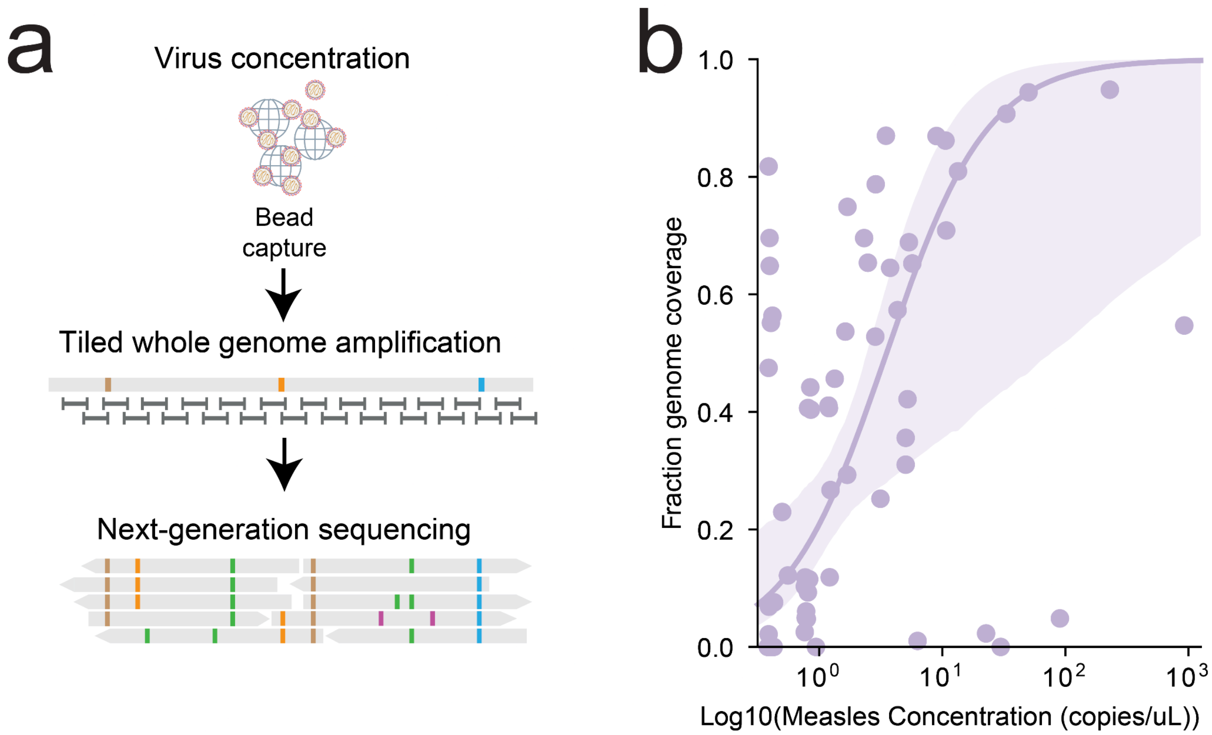


**Supplementary Figure 3: Bead-based virus capture and amplicon sequencing enables near-complete genome coverage a.** Core steps of wastewater processing workflow: virus concentration, whole genome PCR amplification using tiled amplicons, and next generation sequencing. **b.** Genome coverage increases with increasing MeV concentration using Dynabeads virus capture approach. Logistic curve fit (solid line) and 95% bootstrap confidence interval (shaded).

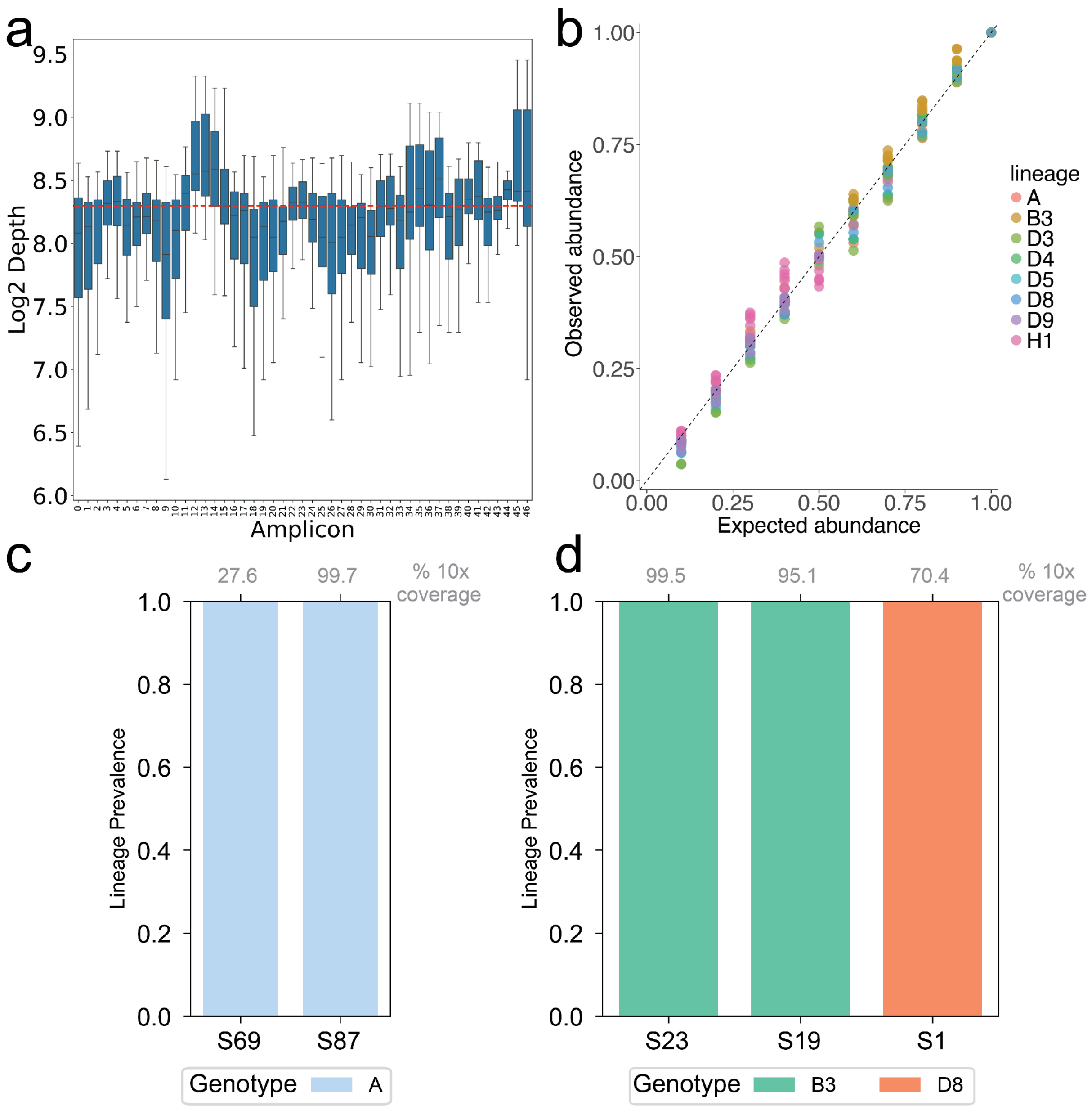


**Supplementary Figure 4: Validation of Freyja bioinformatic approach for MeV using simulation and sequencing of reference samples a.** Boxplots of read depth for each amplicon across all simulated two-component mixtures **b.** Estimated genotype prevalence for using Freyja versus expected prevalence for each component of simulated mixtures **c.** Freyja genotype prevalence deconvolution on vaccine strain control samples. **d.** Deconvolution of sequencing of clinical isolates.

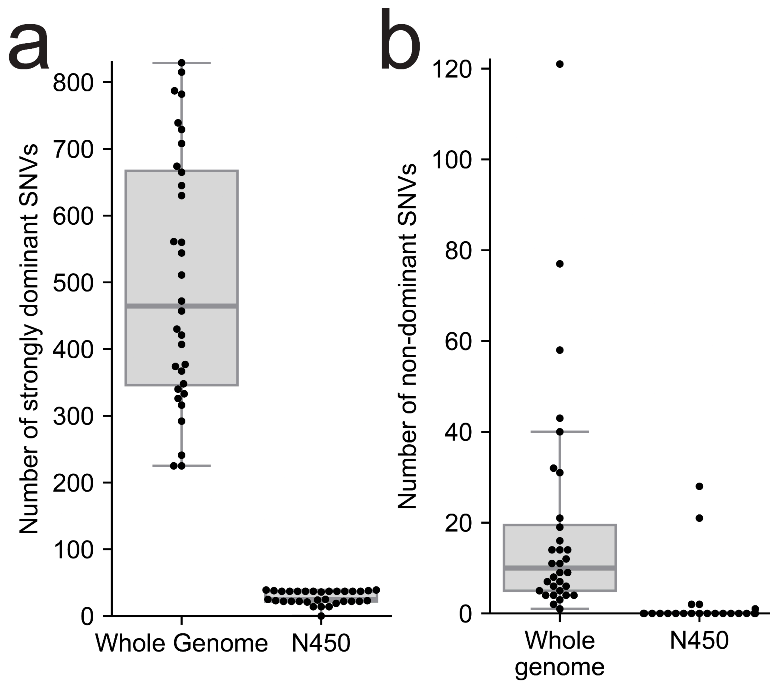


**Supplementary Figure 5: Frequency of single nucleotide variants in wastewater sequencing. a.** Count of mutations with SNV frequency >90%, across the whole genome and just for the N450 region. **b.** Count of non-dominant SNVs with frequency between 10% and 90%.

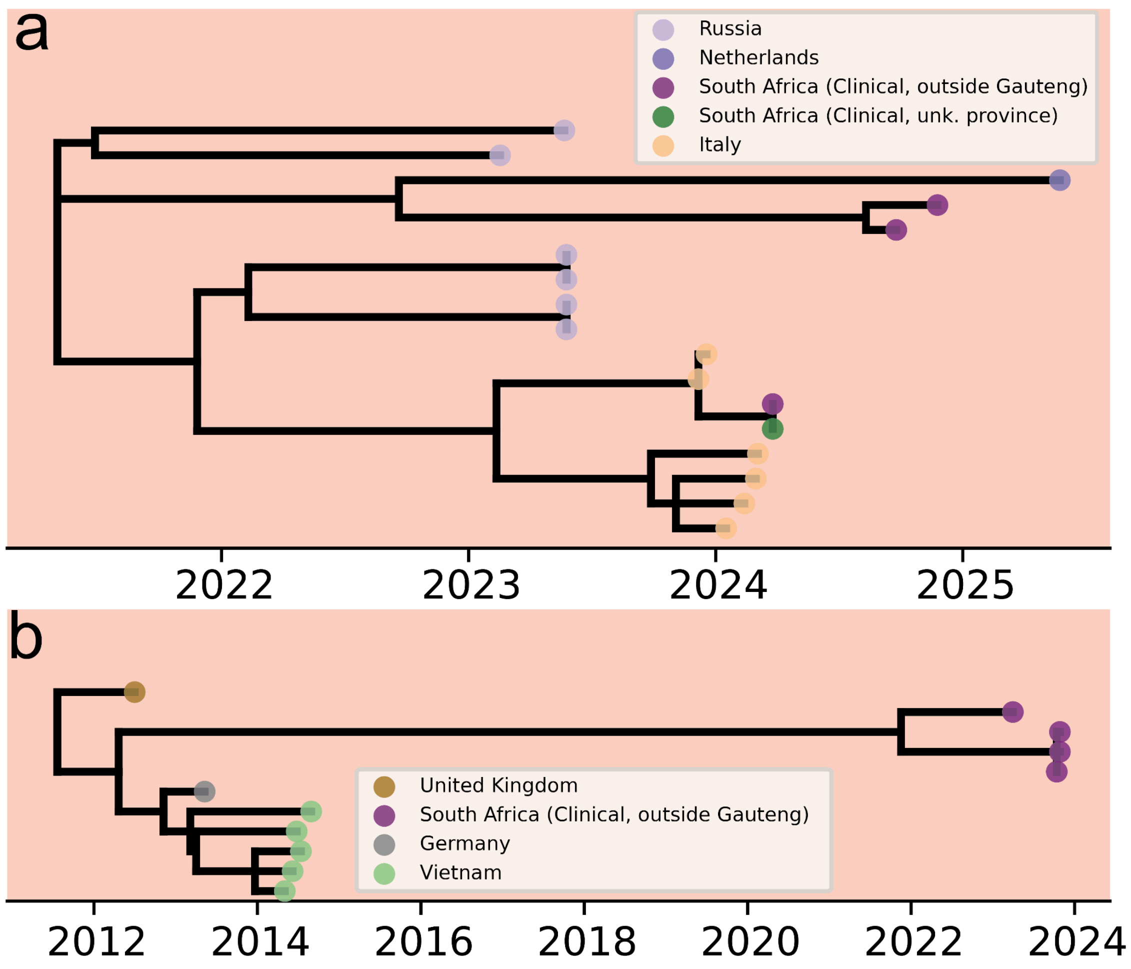


**Supplementary Figure 6: Subtrees for genotype D8 diversity only observed in clinical surveillance a-b.** Time-calibrated subtrees for South Africa-associated clusters from D8 tree in **Figure 4c**, with left cluster (**a**) and right cluster (**b**).

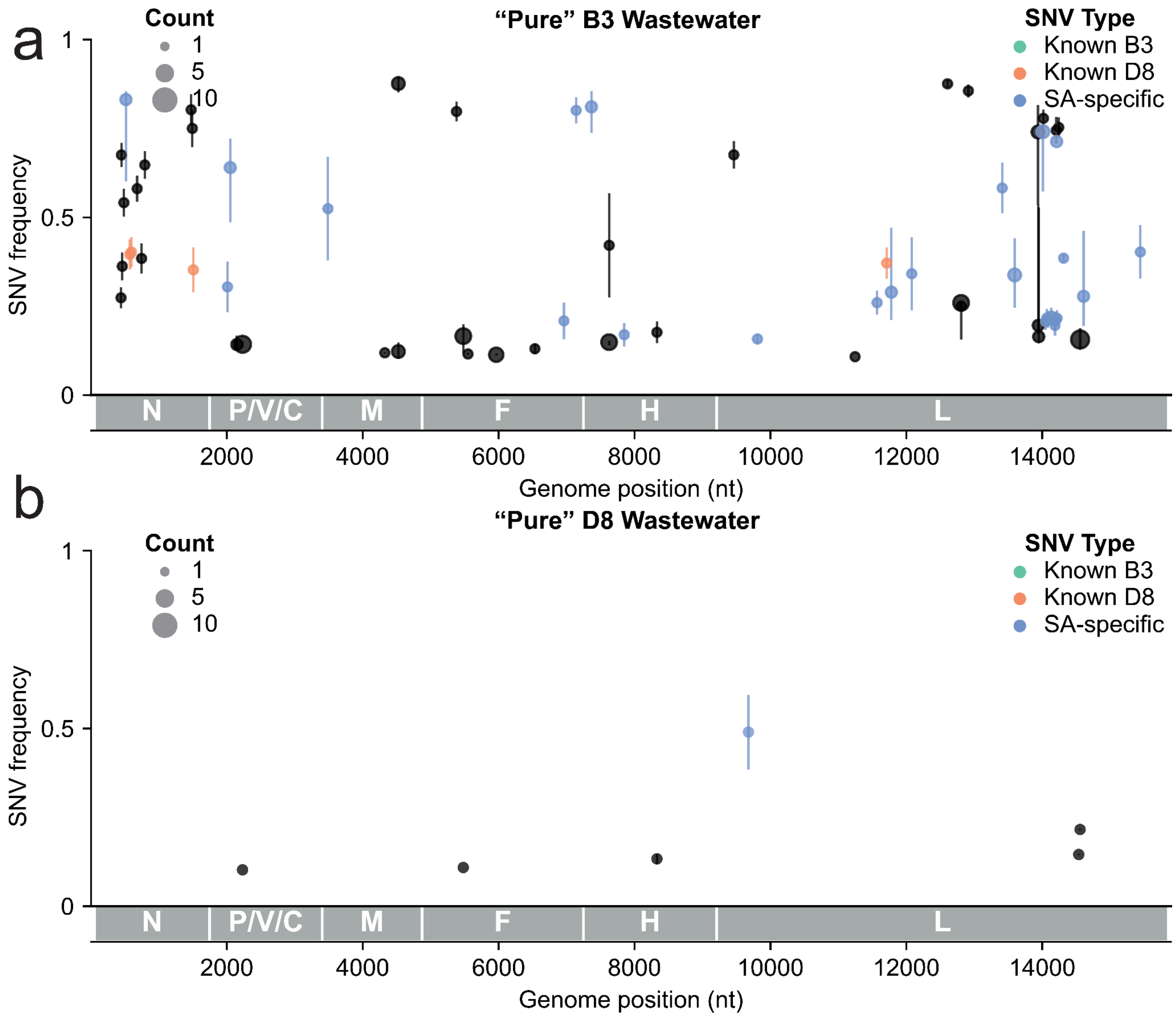


**Supplementary Figure 7: Single nucleotide variant positions and frequency in wastewater. a-b.** Non-dominant SNVs observed >1 time in genotype B3 (**a**) and D8 (**b**) samples over the MeV genome, with median frequency between 10% and 90% when detected. Interquartile range is indicated by a vertical bar. SNVs are colored by genotype association or with known genotype diversity in **Fig. 4a,b**. Genomic regions including nucleoprotein (N), phosphoprotein (P) and accessory proteins C and V, matrix (M), fusion (F), hemagglutinin (H), and large polymerase (L) are indicated by grey bars below the axis.

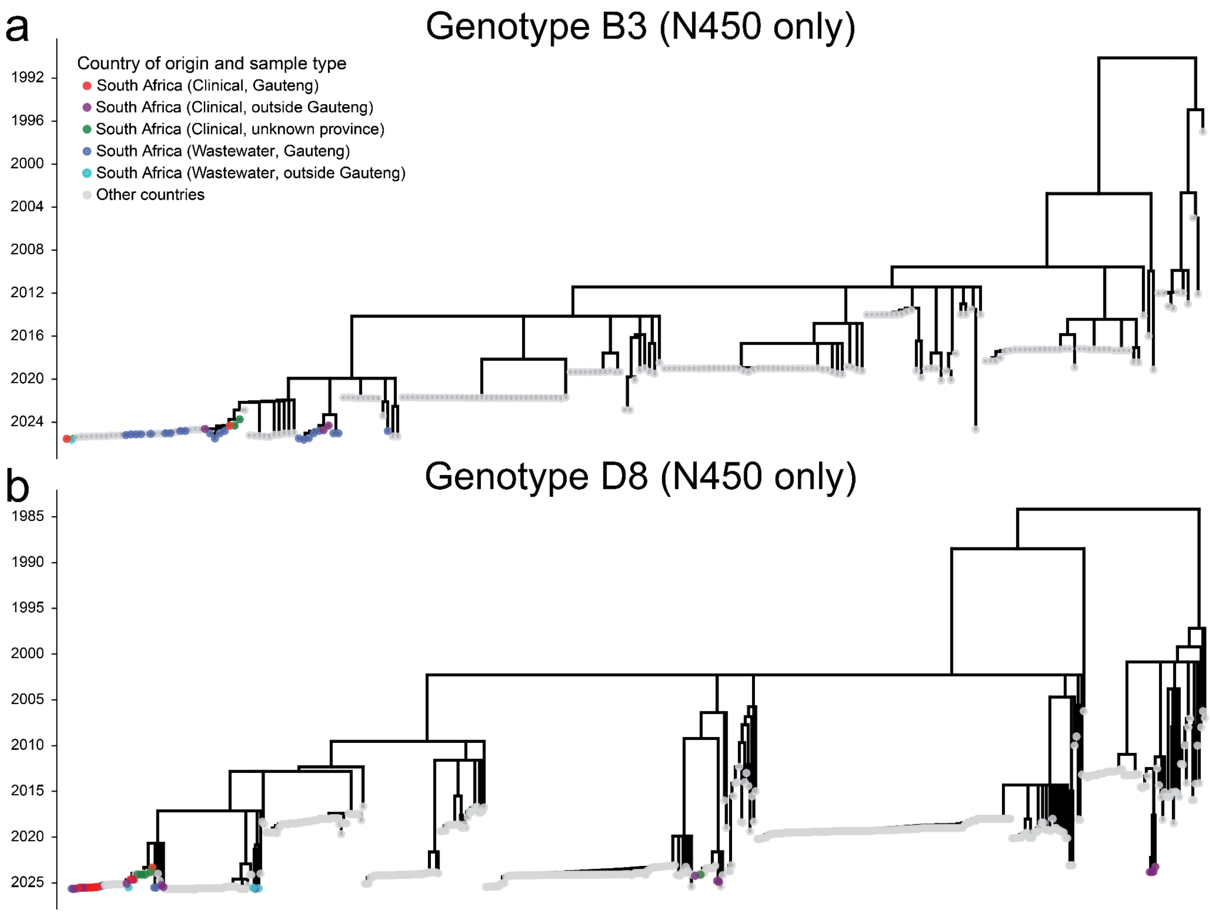


**Supplementary Figure 8: Phylogenetic trees based exclusively on the N450 region. a-b.** Time-calibrated phylogenies using all sequences from **Fig. 4** with N450 region present for genotypes (**a**) B3 and (**b**) D8.
